# Clinical utility of genetic and genomic testing in the precision diagnosis and management of pediatric patients with kidney and urinary tract diseases

**DOI:** 10.1101/2020.04.22.20074690

**Authors:** Nasim Bekheirnia, Kevin E. Glinton, Linda Rossetti, Joshua Manor, Wuyan Chen, Dolores J. Lamb, Michael C. Braun, Mir Reza Bekheirnia

**Author notes:** **Corresponding Author:** Mir Reza Bekheirnia, MD, FACMG, Assistant Professor, Departments of Pediatrics/Molecular and Human Genetics; Baylor College of Medicine, 1102 Bates St., Suite# 245, Houston, TX, 77030. The first two authors contributed equally to this work.

## Abstract

As genetic and genomic testing increasingly integrates into the practice of nephrology, our understanding of the basis of many kidney disorders has exponentially increased. Given this, we recently initiated a Renal Genetics Clinic (RGC) at our large, urban children’s hospital for patients with either a personal or strong family history of renal disorders. Genetic testing was performed in Clinical Laboratory Improvement Amendments (CLIA) certified laboratories using single gene testing, multi-gene panels, chromosomal microarray (CMA), or exome sequencing (ES). A total of 192 patients (185 probands) were evaluated in this clinic, with cystic kidney disease (49/192) being the most common reason for referral followed by Congenital Anomalies of the Kidney and Urinary Tract (CAKUT: 41/192) and hematuria (38/192). Genetic testing was performed for 153 probands with an overall diagnostic yield of 75/153 (49%). In the patients who reached a molecular diagnosis, 13/75 (17.3%), medical or surgical treatment of the patients were impacted, and in 12/75 (16%), previous clinical diagnoses were changed to more accurate molecular diagnoses. Such testing provided an accurate diagnosis for children and in some cases led to further diagnosis in seemingly asymptomatic family members and changes to overall medical management. Molecular testing, as facilitated by such a specialized clinical setting, thus appears to have clear utility in the diagnosis and counseling of patients with a wide range of renal manifestations.

## Introduction

Genetic and genomic testing has increasingly integrated into the practice of different specialties in medicine and surgery. Within the field of nephrology in particular, the availability of such testing led to the rapid growth and expansion of our knowledge of the clinical spectrum of genetically-mediated renal diseases. The molecular etiology of Mendelian genetic renal diseases, such as polycystic kidney disease, Alport syndrome, several forms of monogenic steroid-resistant nephrotic syndrome (SRNS), and nephronophthisis has grown and can now be identified in a significant portion of affected individuals. In patients with SRNS, 29.5% of those diagnosed before age 25 will have a pathogenic variant in one of 30 known SRNS genes.^1^ Even in a condition not commonly associated with genetic causes, such as nephrolithiasis, around 15% of individuals have a specific underlying genetic etiology.^2^ Given the growing number of recognized disease-causing gene defects, next-generation multi-gene panels are now available and in some cases, can provide adequate diagnostic coverage.^3^ Similarly, exome sequencing (ES) has immense utility in the diagnosis of adults and children with a variety of disorders.^4,5^

With the expanding number of candidate genes and the increasing complexity of genetic testing available, the need for more comprehensive diagnostic evaluations for such patients increased, as well. To address this need, a Renal Genetics Clinic (RGC) at Texas Children’s Hospital (TCH) was formed in February of 2015. Patients are referred from a variety of care settings including the Pediatric Nephrology Clinic and various inpatient/outpatient services at TCH. Through this clinic, patients undergo a thorough genetic evaluation with a focus on kidney-specific malformations, complications or diseases. Furthermore, given the nature of the clinic, family members of affected individuals can be evaluated allowing us to provide guidance, if needed, for family planning. Extensive research shows the key roles genetic and genomic defects play in pediatric renal disorders, and growing number of studies are evaluating the utility of clinical genetics evaluation and genetic testing in the clinical practice.^6,7^ Therefor there is still a need to expand the knowledge in the intersection of clinical nephrology and clinical genetics. The specific objective of this study is to assess the role of clinical genetics in precision diagnosis and management of early-onset pediatric renal diseases. Diagnostic yield and impact on medical management is reported for the first four years of this clinic’s operations.

## Methods

Study participants: Patients were all evaluated within the RGC at TCH. The clinic was initially held on only one half-day per month, though this was increased to a full clinical day monthly after approximately 18 months. Patients were interviewed and examined by a clinical geneticist and appropriate genetic testing was recommended based upon their clinical history, presentation and family history. Pretest counseling was provided. Patients consented for ES based on the performing CLIA laboratories’ consent forms that include secondary findings (medically actionable, and carrier status). The genetic variants reported in this study were classified only by CLIA laboratories. Reported variants by CLIA laboratories were evaluated by a clinical geneticist during clinical visits aiming to provide a clinical diagnosis and to discuss pertinent management. Clinical information on subjects was collected retrospectively for the period of February 2015 to June 2019. All patients seen during this timeframe were eligible to participate. Institutional Review Board (IRB) approval was obtained to perform a retrospective cross-sectional to study the yield and impact of genetics evaluation. Outcome of the study was defined as the “impact” of genetic evaluation on diagnosis and management of patients and were classified in six categories: 1) impact on medical and/or surgical treatment, 2) change of medical diagnosis, 3) providing diagnostic certainty, 4) subsequent evaluation for other body system involvement, 5) cascade family member testing, 6) molecular confirmation. To minimize the bias regarding scoring of the outcome, a pediatric nephrologist blinded to the identifiers reviewed the allocated scores.

### Genetic Testing

Testing performed by CLIA laboratories include: disease specific panels, chromosomal microarray (CMA), expanded next generation sequencing panel (Total Blueprint^®^), and ES [trio or proband-only (when both parents were not available)]. When appropriate, combinations of these tests were also performed to optimize diagnostic yield in cases with atypical or unclear phenotypes. Comparison analysis of detection rate between different testing modalities was not performed because Genomic DNA was isolated from peripheral leukocytes obtained via venipuncture or less commonly from saliva.

## Results

A total of 192 patients (185 probands) were evaluated in this clinic over the period of time from 02/2015 – 06/2019 (Table 1). Patients ranged in age from 1 day of life to 25 years of age with a mean age of 8.7 years (SD 6.0). The most common reason for referral was cystic kidney disease in 49 patients (47 probands) (25.5%) followed by CAKUT in 41 patients (21.3%), 38 patients (36 probands) with hematuria (19.8 %), and 21 patients (19 probands) with proteinuria (10.9%). A further 43 patients (42 probands) (22.5%) were seen for “other” clinical diagnoses including nephronophthisis, nephrocalcinosis, developmental delay combined with renal disease or overlapping phenotypes (Supplemental Table 1). From 185 probands, four were asymptomatic and genetic testing was only recommended for their affected parents, however, such testing has not been completed to date. Genetic testing was performed for 153 of 181 probands (84.5%). We were not able to perform genetic testing for 28 individuals (14 because of insurance denial, and 14 families were not interested to pursue genetic testing). Among 153 probands, 75 (49%) had positive diagnostic results (Table 2). In an additional four patients, the patients’ phenotype was partially explained based upon the genetic work up (Supplemental Table 2).

**Table 1.** Demographics and indications for referrals among 192 patients evaluated at Renal Genetics Clinic between 2/2015-6/2019.

| **Gender:** |  |
| --- | --- |
| Male (%) | 102 (53.1) |
| Female (%) | 90 (46.9) |
| **Total Patients Evaluated:** | **192** |
| **Ethnicity:** |  |
| African American (%) | 29 (15.1) |
| Asian (%) | 1 (0.1) |
| Caucasian (%) | 67 (34.9) |
| Latino (%) | 67 (34.9) |
| Mixed (%) | 25 (13) |
| Other (%) | 3 (1.6) |
| **Age at Evaluation:** |  |
| Mean (Std Deviation) | 8.7 years (6.0) |
| Range | 1 day - 25 years |
| **Initial Indications for Referral:** |  |
| CAKUT (%) | 41 (21.3) |
| Cystic Kidney Disease (%) | 49 (25.5) |
| Hematuria (%) | 38 (19.8) |
| Proteinuria (%) | 21 (10.9) |
| Other (%) | 43 (22.5) |

**Table 2.**
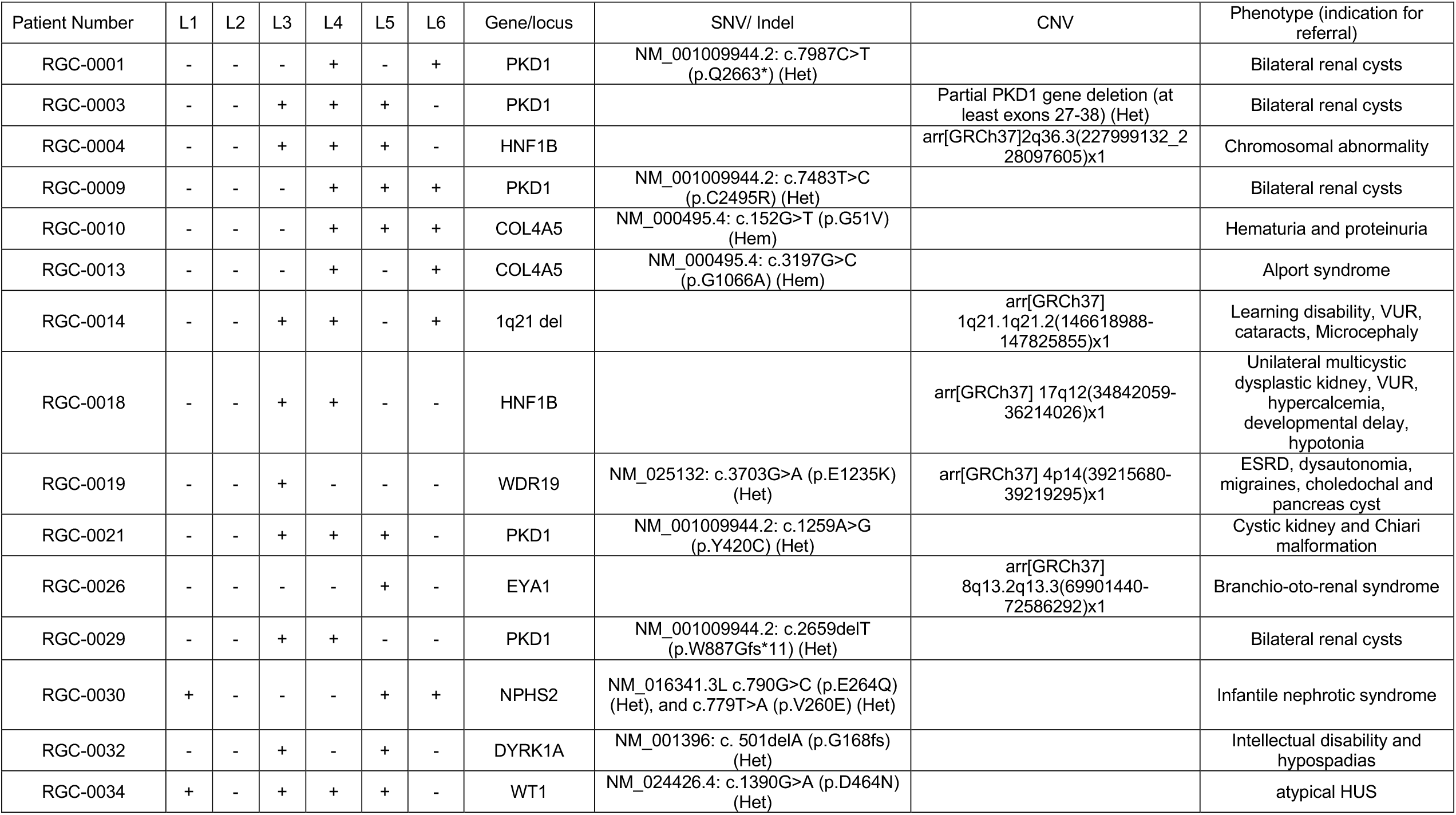
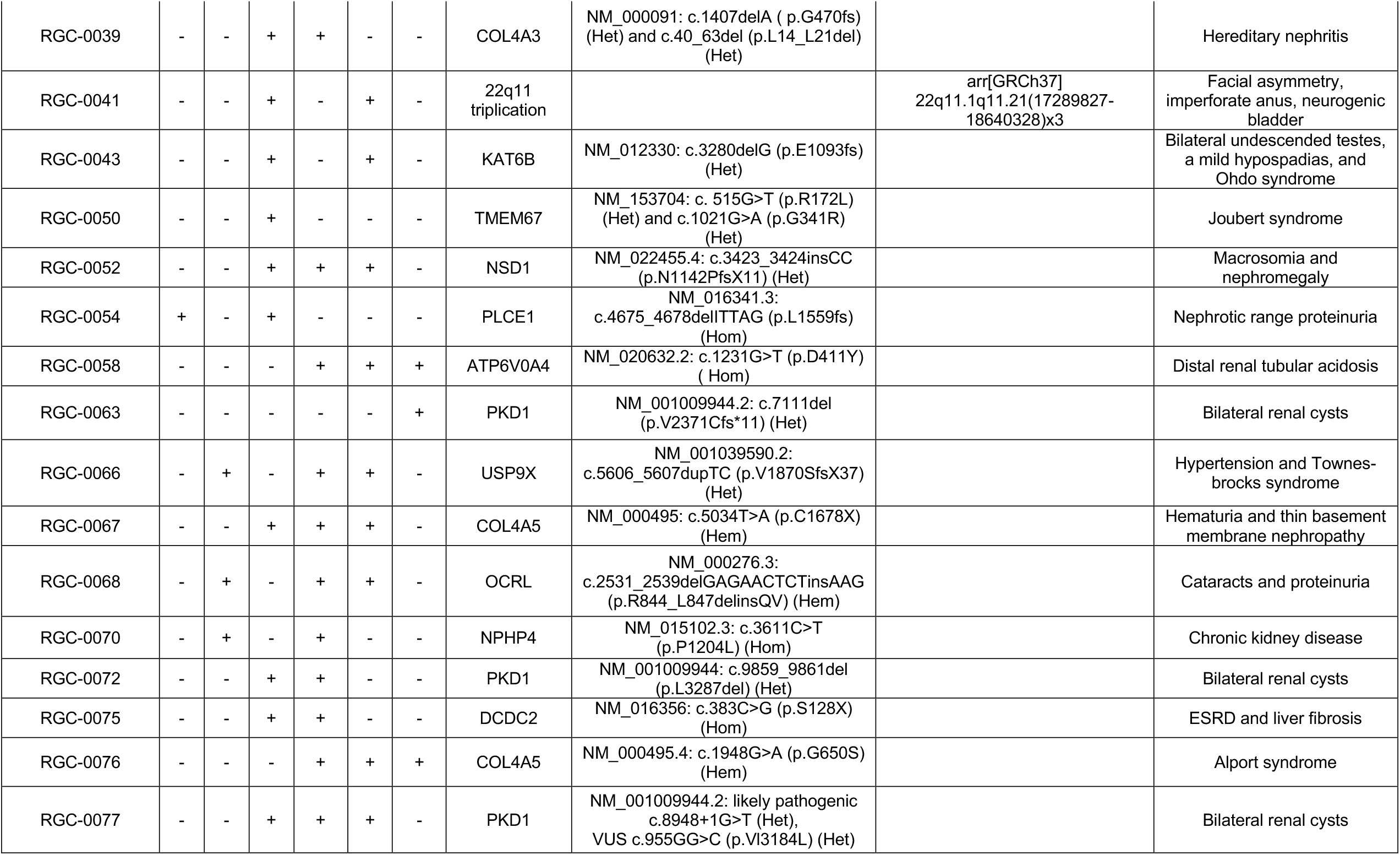
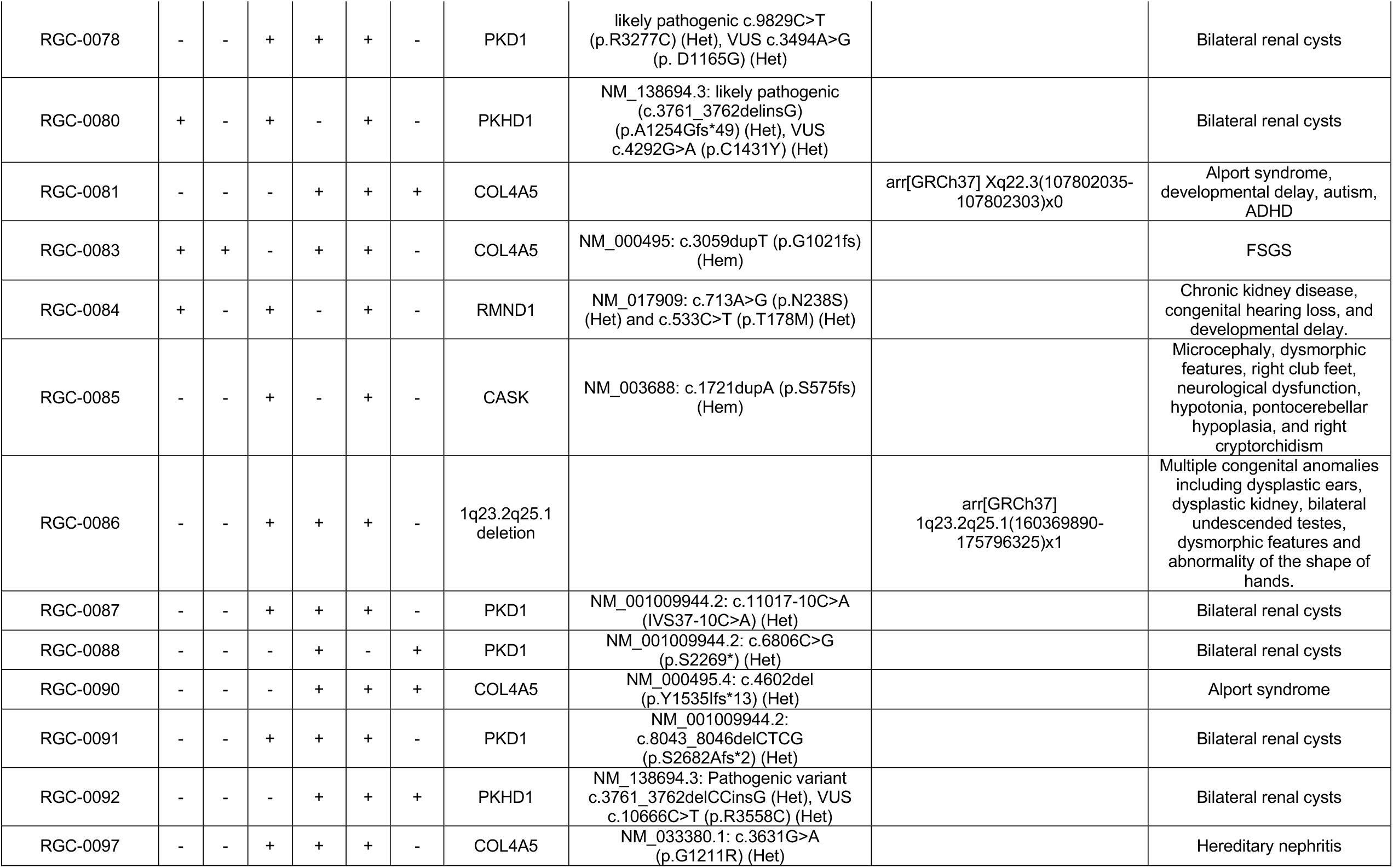
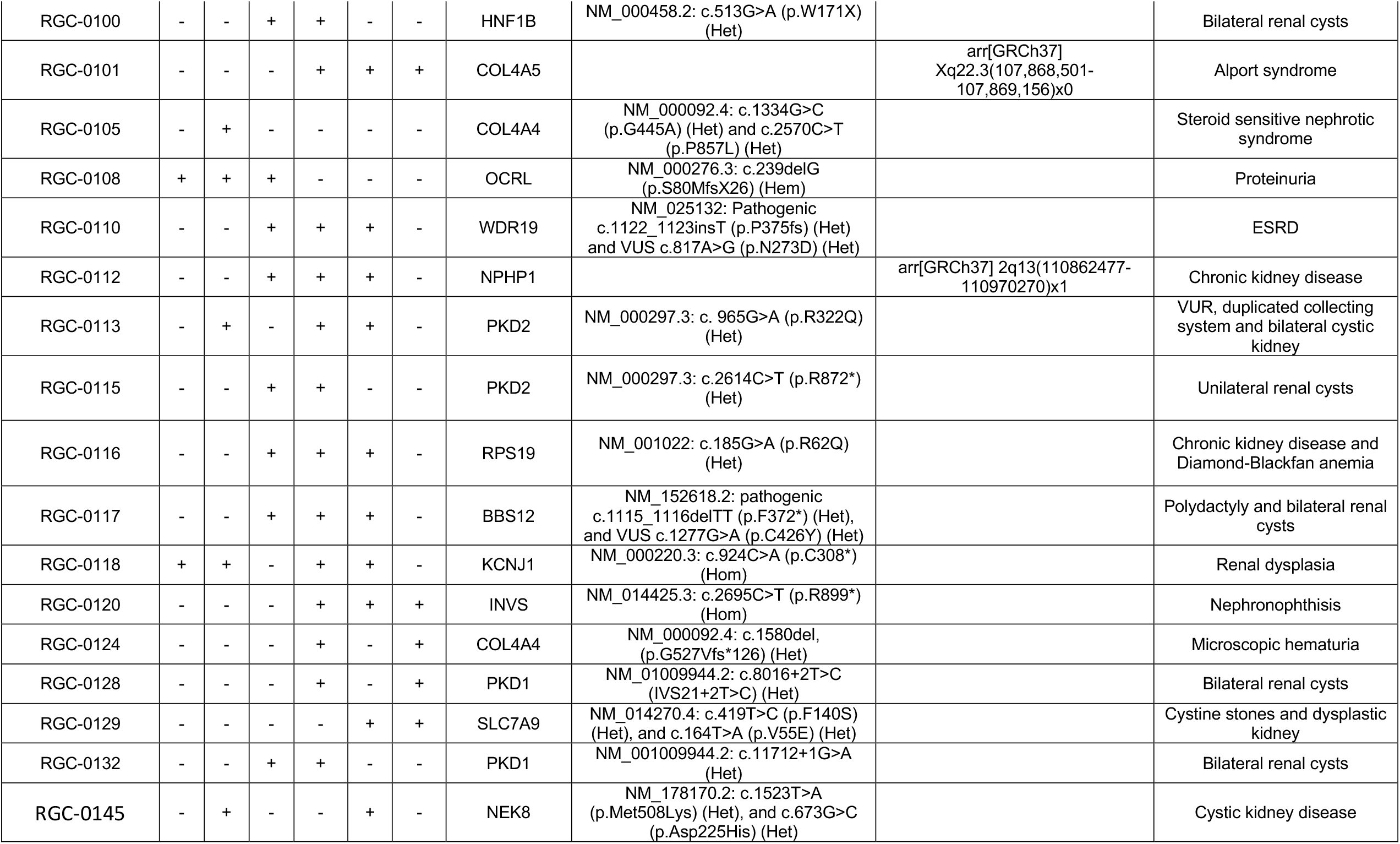
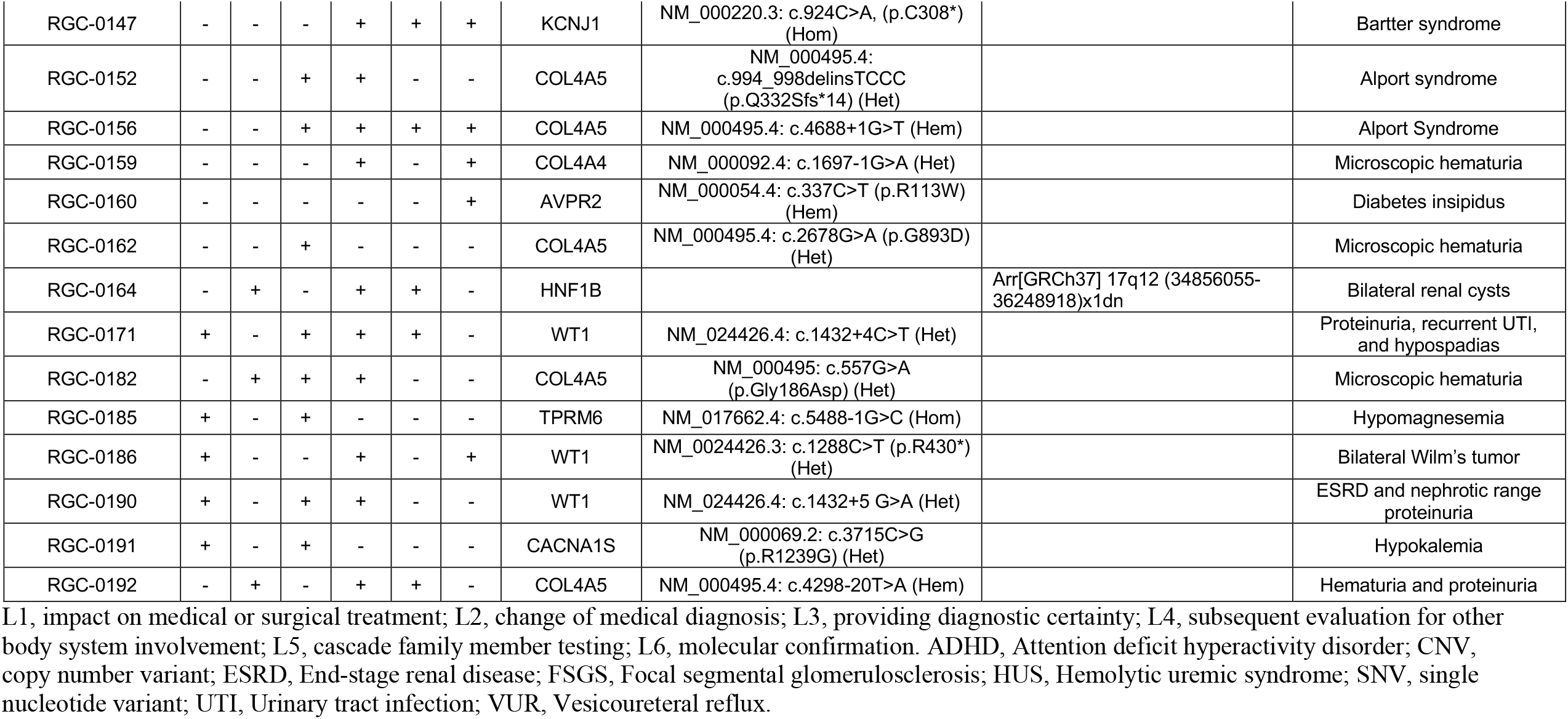
Impact of genetic and genomic testing and evaluation on medical management of 75 patients (referred to Renal Genetics Clinic) with positive diagnostic results.

Given the breadth of diagnoses encountered, no single test was universally applicable to every patient. Different tests, or combination of tests, were recommended and completed for patients depending on the specificity of their clinical phenotype or reported history through different CLIA laboratories. Supplemental Table 3 summarizes the detection rate of different tests among indications for referral. For patients with CAKUT (41 probands; tests completed for 33), for instance, CMA or a combination of CMA with ES led ultimately to a diagnostic yield of 42.4% (14/33) (including the partially diagnosed cases). However, in patients who presented with hematuria, the use of a multi-gene panel was the most successful approach to provide a molecular diagnosis in 77% (10/13) cases. Patients with hematuria were in fact the most likely to receive a molecular diagnosis with an overall diagnostic yield of 66.7% (20/30) when a combined approach of a targeted panel, CMA and ES was utilized.

Our testing approach led to the identification of pathogenic or likely pathogenic single nucleotide variants (SNVs) in 31 genes (Table 2). Similarly, 13 different pathogenic or likely pathogenic copy number variants (CNVs) were also identified, ranging from single exon deletions to large megabase-sized deletions of multiple genes. Pathogenic SNVs or CNVs were found most commonly in *PKD1* (14) followed by *COL4A5* (11), *HNF1B* (3), *COL4A4* (3) and *PKHD1* (3).

### Impact on precision diagnosis and management

To assess the impact of genetic testing and evaluation on patients’ management, each patient with a positive result was scored according to a 6-level scoring system as detailed in the Methods. Out of 75 positive diagnostic results, 13 (17.3%) affected immediate medial or surgical treatment (L1, Table 2). The most common referral indication among these 13 patients was nephrotic syndrome or proteinuria, a condition where medication adjustment by avoiding immunosuppression became possible. Other immediate benefits of genetic evaluation included surgical decision-making regarding the need for prophylactic (RGC-0034) or therapeutic nephrectomy (RGC-0186) in patients with pathogenic *WT1* variants. In three patients (RGC-0118, RGC-0185, and RGC-0191), targeted treatment recommendations with directed pharmacotherapy (indomethacin, magnesium, and acetazolamide) became possible after identification of underlying molecular diagnosis (*KCNJ1, TPRM6, and CACNA1S*).

Accurate molecular diagnoses were achieved in 75 patients and among them: (1) twelve (16%) prior diagnoses were changed including Four changes from FSGS to Alport syndrome; (2) in 43, diagnostic certainty became possible only with genetic testing; and (3) in 23, molecular confirmation of an already existing clinical diagnosis was provided. Other impacts on management included evaluation of other body organ systems and cascade family testing in 55 and 45 patients, respectively. In three families, reproductive genetic counseling immediately affected the family’s decision-making for their family planning. Figure 1 summarizes the number of patients in each of the six levels of our scoring system. Other important impacts on management included screening of potential living related kidney donors, planning for solid organ transplantation, and accurate genetic counseling. The discovery of inherited pathogenic variants in autosomal dominant disease genes led, for instance, to the discovery of previously unrecognized clinical abnormalities in parents (e.g. RGC-0003 and RGC-0087) and the illumination of unusual inheritance patterns (e.g. pseudodominance, RGC-0080)

**Figure 1.**
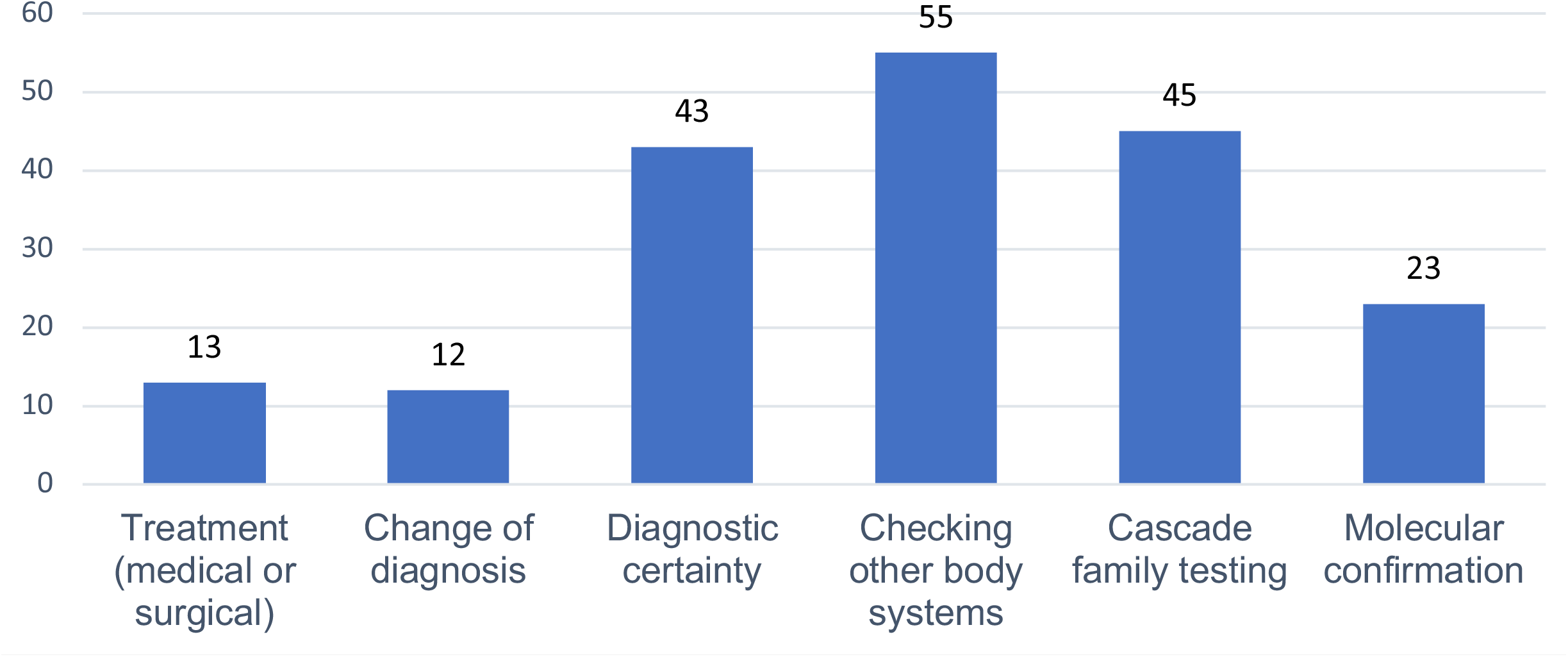
Number of probands impacted by each of the 6-level scoring system

## Discussion

In this study, the detection rate (75/153, 49%) and the clinical utility of genetic evaluation/testing was demonstrated for pediatric renal disorders in a Renal Genetics Clinic setting. In 29.3% (22/75) of the patients with positive results, immediate medical/surgical treatment was impacted or the prior diagnoses (achieved by either biopsy or clinical evaluation) were changed.

RGCs are the optimal mechanism for integrating a comprehensive genetic evaluation with appropriate molecular testing on a clinical basis.^8^ This kind of clinic also allows for a family centered approach where unaffected relatives may also be evaluated and counseled on their risks for renal disease. Examples of these clinics in Australia, the UK, and China, showed diagnostic yields of 46%,^9^ 42%,^10^ and 42.1%^7^ respectively. Our diagnosis yield is at the same scale (40-50%), but slightly higher which could be explained by following factors. First, the indications of referral among these different clinic models are not the same. Second, broad genetic testing options were available in our center. Lastly, patients were referred with rigorous initial evaluation by pediatric nephrologists. A less stringent referral criteria may lead to larger number of patients seen with a higher number of total positive diagnosis, however with overall lower diagnosis rate.

Our testing approach utilized various combinations of targeted panels, CMA and ES (by CLIA laboratories). This resulted in the identification of pathogenic SNVs in 31 different genes and 13 unique pathogenic CNVs. Many of these changes are novel and have not previously been reported in published databases. In terms of testing performance, our diagnostic yield is higher than the reported yield of ES for adult patients with kidney disease,^11^ highlighting the increased contribution of genetic abnormalities in the pediatric population. The diagnostic rate of CAKUT in this cohort is higher than expected from literature.^12^ This is likely due to stringent referral criteria that select syndromic patients.

Our patients were placed into one of five categories based upon their clinical presentation and presumed diagnosis. Each category varied in terms of which molecular testing was felt to be most appropriate both initially and upon follow-up. For instance, panel testing (known to be cost-effective and specific) was very useful in cases of both cystic kidney disease and hematuria. For patients with cystic kidneys in particular, a panel appeared to be a good initial diagnostic choice because of the high prevalence of *PKD1* pathogenic variants. If this test result was negative, or if patients had other concerning physical or clinical abnormalities, expanded testing could be pursued with ES or CMA. This allowed us to identify diagnostic variants in genes not previously considered. For instance, a patient initially referred for cystic renal disease was later found to have a pathogenic variant in *HNF1B* more commonly associated with CAKUT (RGC-0164) while another patient was diagnosed with biallelic variants in *BBS12* indicative of Bardet-Biedl syndrome (RGC-0117).

Most importantly, genetic evaluation resulted in recommendations for immediate medical or surgical treatment in 17.3% of probands. In addition, the original diagnosis in 16% of probands was changed. The benefits of level 1 impact on management included targeted therapies, and preventing the use of inappropriate treatments (i.e. corticosteroids where there was no expectation of benefit). We compared diagnosis pre- and post-genetic evaluation, and concluded that genetic testing improved diagnostic accuracy given that the diagnosis might be different from what was previously achieved by clinical or pathological evaluations. The change of diagnosis from FSGS to Alport, reported in this study, was also published by other investigators.^13^ Additional benefits included reducing the use of invasive diagnostic procedures, such as renal biopsy. Reduction of genomic testing costs will ultimately result in the precise diagnosis of patients for whom an initial syndromic diagnosis was not clinically suspected. In addition to confirmatory diagnosis, a molecular diagnosis may also provide prognostic information, establish a targeted surveillance for extrarenal complications and facilitate renal transplant and reproductive planning.^6^

This study however has the following limitations. First, the design of this study is retrospective and there is still a need for larger, prospective studies moving forward. Second, patients’ viewpoints of genetic or genomic testing was not studied. Third, though our study included a range of diagnoses, the relatively small overall number/type of patients evaluated in this clinic may affect generalization of our data. Fourth, only pediatric population was studied. Finally, while we have investigated the health impacts of genetic testing, the economic impact of this testing in kidney disease was not studied.

Strengths of this study include: (1) the ability to perform advanced clinical genetic testing for a large proportion of our patients; (2) the diversity of the cohort; specifically their ethnicity, renal phenotypes, clinical diagnoses and severity of kidney disease; (3) access to world-class pediatric nephrology and clinical genetics groups; and (4) affiliation with one of the largest children’s hospitals in the United States.

## Data Availability

N/A

## Acknowledgements

The authors would like to acknowledge the many patients and their family members. We would also like to thank the following diagnostic labs for their superb genetic/genomic testing: Baylor Genetics, GeneDx and Prevention Genetics. The authors would additionally like to thank all referring physicians from either within the Texas Children’s Hospital medical system or outside. This study was supported in part by the Multidisciplinary K12 Urologic Research (KURe) Career Development Program K12DK0083014 from the National Institute of Digestive Diseases and Kidney to DJL (MRB was a K12 scholar), R01DK078121 from the National Institute of Digestive Diseases and Kidney to DJL. DJL is also supported in part by the Frederick J. and Theresa Dow Wallace Fund of the New York Community Trust. This study was also supported by start-up funds from Renal section, Department of Pediatrics at TCH and BCM.

